# Quantifying treatment effects of hydroxychloroquine and azithromycin for COVID-19: a secondary analysis of an open label non-randomized clinical trial

**DOI:** 10.1101/2020.03.22.20040949

**Authors:** Andrew A. Lover

**Author notes:** https://www.umass.edu/sphhs/person/faculty/andrew-lover. The author stands by all analytical and statistical aspects of this preprint. However, subsequent to this analysis, further details of the original study have been released-with major uncertainties in study design, reporting, choice of endpoints, and most importantly, data integrity [1, 2]. Therefore, all results from the original study should be viewed with considerable skepticism.

## Abstract

Human infections with a novel coronavirus (SARS-CoV-2) were first identified via syndromic surveillance in December of 2019 in Wuhan China. Since identification, infections (coronavirus disease-2019; COVID-19) caused by this novel pathogen have spread globally, with more than 250,000 confirmed cases as of March 21, 2020. An open-label clinical trial has just concluded, suggesting improved resolution of viremia with use of two existing therapies: hydroxychloroquine (HCQ) as monotherapy, and in combination with azithromycin (HCQ-AZ). [3, 4].

The results of this important trial have major implications for global policy in the rapid scale-up and response to this pandemic. The authors present results with p-values for differences in proportions between the study arms, but their analysis is not able to provide effect size estimates.

To address this gap, more modern analytical methods including survival models, have been applied to these data, and show modest to no impact of HCQ treatment, with more significant effects from the HCQ-AZ combination, potentially suggesting a role for co-infections in COVID-19 pathogenesis.

The trial of Gautret and colleagues, with consideration of the effect sizes, and p-values from multiple models, does not provide sufficient evidence to support wide-scale rollout of HCQ monotherapy for the treatment of COVID-19; larger randomized studies should be considered. These data also suggest further randomized-controlled studies of HCQ-AZ combination therapy should be undertaken.

## 1 Introduction

Evidence-based public health programming is essential for global pandemic planning, and optimization of resources. However, unadjusted analyses may provide distorted estimates, or not full utilize scarce clinical data, especially in with consideration of intention-to-treat analyses, where all persons enrolled are analysed [5, 6]. Finally, effect size estimates allow for the magnitude of potential clinical impact to considered in policy adoption, as statistical significance may not be biologically important [7].

## 2 Data and Analysis

All analyses was performed using Stata 16.1 (College Station, TX, USA). Standard 95% confidence interval were used; model parsimony was assessed using Akaike and Bayesian information criteria (AIC/BIC). All models were adjusted for age and sex. Assessing the predictive power of logistic models used Tjur’s 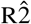. [8].

### 2.1 Data source

Data were obtained from [4], and the Supplemental Table 1 was digitized using Tabula software, with subsequent hand-validation. Data for the six patients who were lost-to-follow-up (LTF) were manually entered into a database.

**Table 1:** Risk ratios for clearance of virema, by day six, using binary regression (Primary outcome). (N=30).

| variable | RR | 95% CI | p-value |
| --- | --- | --- | --- |
| **Study Arm** |  |  |  |
| Control | reference | - | - |
| HCQ | 3.836 | 1.020 - 14.42 | 0.047 |
| **Age** Years | 1.009 | 0.996 - 1.022 | 0.176 |
| **Sex** |  |  |  |
| Male | reference | - | - |
| Female | 0.585 | 0.335 - 0.963 | 0.176 |

**Table 2:**
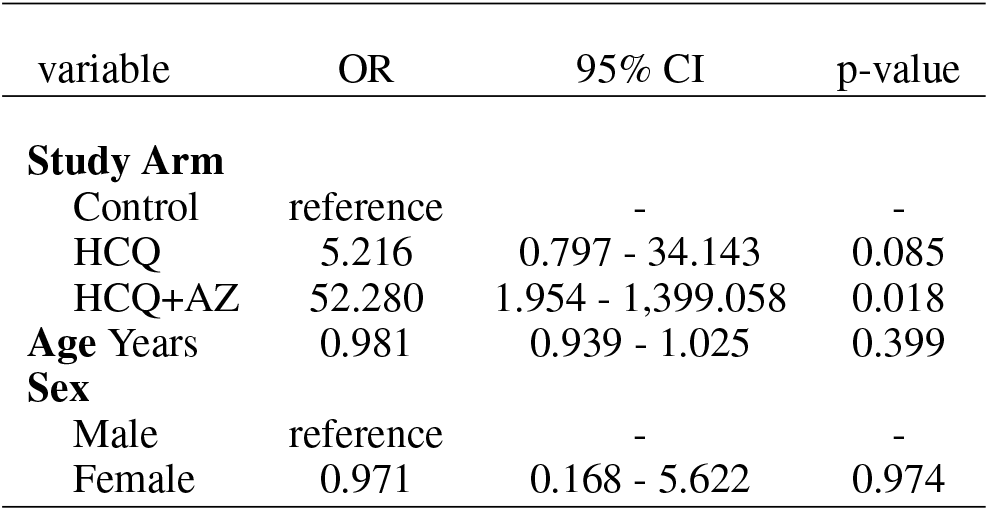
Odds ratios for clearance of viremia, by day six, from a Firth penalized likelihood regression (Primary outcome). (N=30).

| variable | OR | 95% CI | p-value |
| --- | --- | --- | --- |
| **Study Arm** |  |  |  |
| Control | reference | - | - |
| HCQ | 5.216 | 0.797 - 34.143 | 0.085 |
| HCQ+AZ | 52.280 | 1.954 - 1,399.058 | 0.018 |
| Age Years | 0.981 | 0.939 - 1.025 | 0.399 |
| **Sex** |  |  |  |
| Male | reference | - | - |
| Female | 0.971 | 0.168 - 5.622 | 0.974 |

**Table 3:** Hazard ratios for time-to-first negative PCR, using flexible parametric models. (N=36). (Secondary Outcome)

| variable | HR | 95% CI | p-value |
| --- | --- | --- | --- |
| **Study Arm** |  |  |  |
| Control | reference | - | - |
| HCQ | 3.026 | 0.925 - 9.895 | 0.067 |
| HCQ+AZ | 7.073 | 1.722 - 29.044 | 0.007 |
| Age Years | 0.982 | 0.955 - 1.010 | 0.213 |
| **Sex** |  |  |  |
| Male | reference | - | - |
| Female | 1.016 | 0.365 - 2.825 | 0.976 |

### 2.2 Primary outcome

The primary outcome as reported by the authors “The primary endpoint was virological clearance at day-6 post-inclusion.” An optimal analysis for this endpoints in a binary regression which avoids many of the potential biases in logistic models when outcomes are common [9].

### 2.3 Secondary outcome

The stated “Secondary endpoint was virological clearance overtime during the study period.” and standard Cox survival time models and Royston-Parmar flexible parameteric models [10] were used to capture the time-to-first negative PCR (with a Ct threshold of >=35). The incorporation of the censored patients was as per standard methods.

## 3 Results

The sequence of confirmed viremia via PCR is shown in (Fig. 1), and the LFT patients are at the bottom of the figure. The primary outcome was assessed using binary regressions to provide relative risks for clearance of viremia between the study arms. The main effect of interest (that of all combined HCQ-treated patients versus control), shows a marginally significant risk ratio of 3.84 (95 % CI 1.02 - 14.42, p= 0.047). Analysis of the separate HCQ and HCQ+AZ outcome was not possible due to quasi-separation of the model.

**Figure 1:**
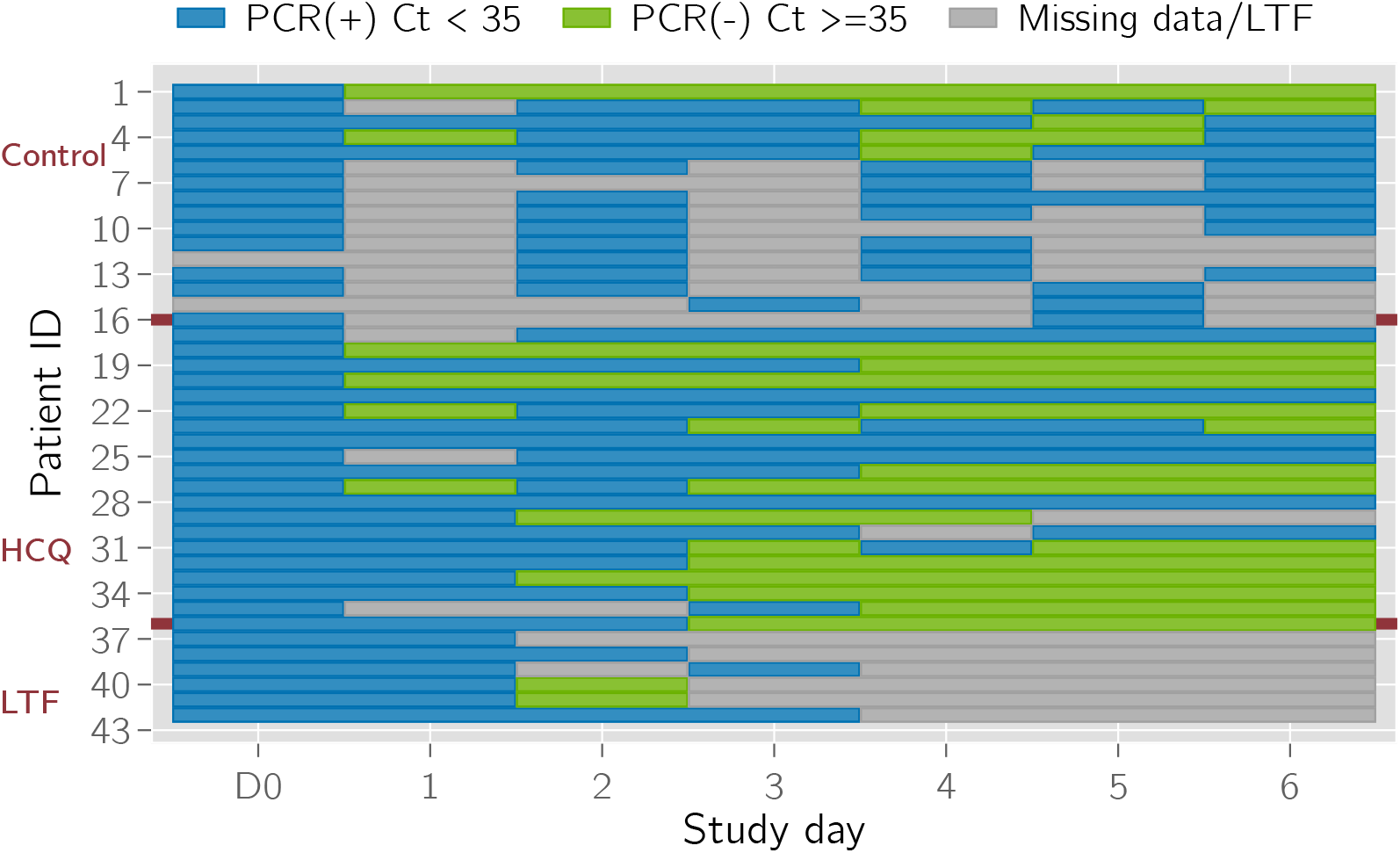
Sequence plot of enrolled patients. (N= 42).

**Figure 2:**
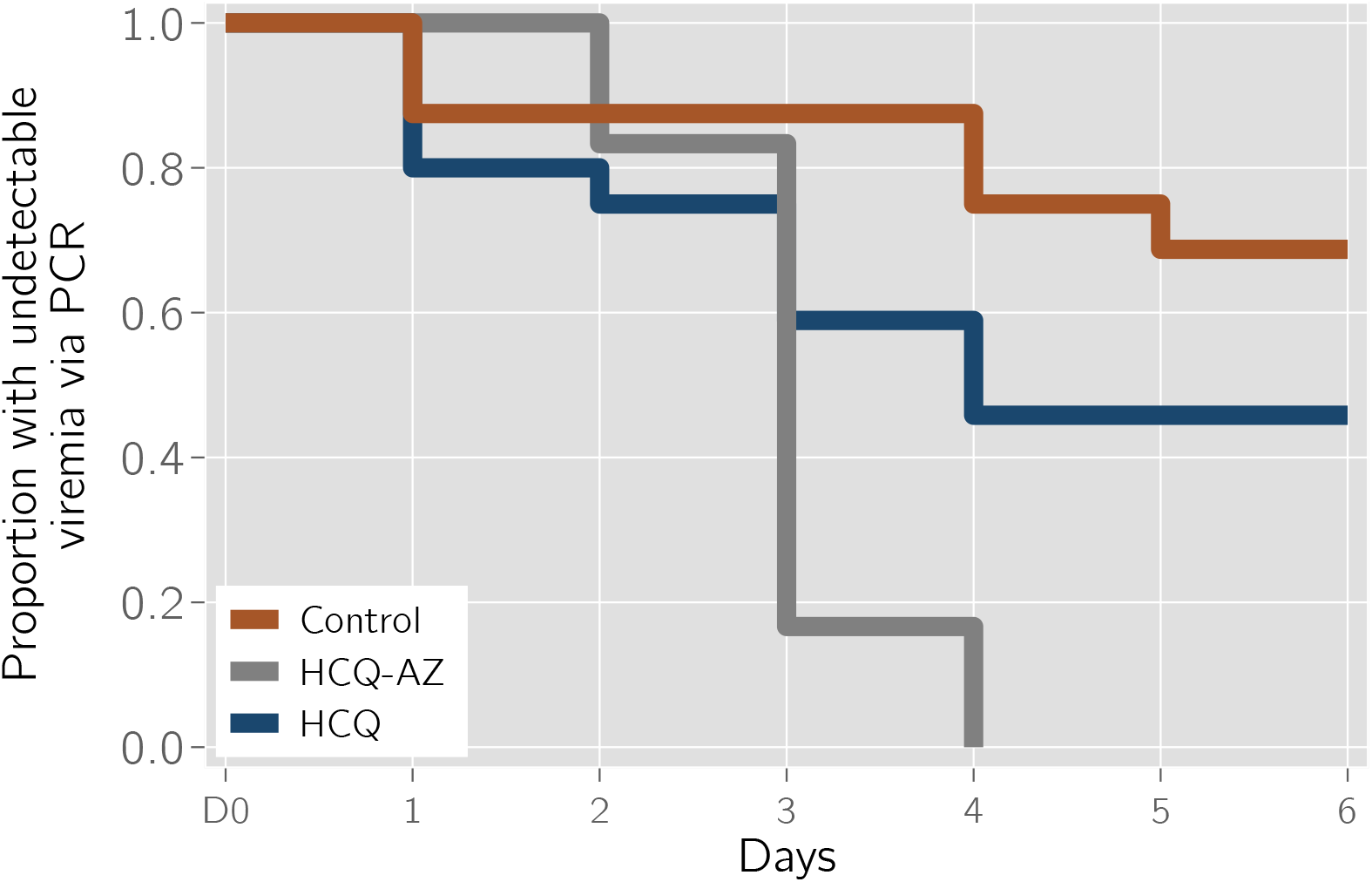
Unadjusted Kaplan-Meier plot of all enrolled patients (N= 42).

To address the limitations of these models, Firth penalized-likelihood model were used, which deal well with separation and quasi-separation. [11]

To assess the secondary outcome, Kaplan-Meier and Cox models were use initially to compare the time-to-event with adjustment for covariates. Due severe proportional hazard violations in the HCQ-AZ group, flexible parametric models were used to estimate effects.

## 4 Discussion and Conclusions

Together these results, especially in consideration of the loss to followup of six patients, do not provide sufficient evidence to support HCQ monotherapy for the treatment of COVID-19.

This analysis is not without limitations: interpretation of the presented dataset may be incorrect; the analysis models for the primary and secondary outcome endpoints is inferred from the original authors’ description and power calcualtions (case-control design); and not all covariates were available in the original data [12].

However, taken together, this analysis does suggest further studies of HCQ-AZ combination therapy should be prioritized with great haste. The rapid increase in confirmed infections within the last few days suggests that the pandemic is accelerating, and there are major opportunity costs associated with all choices [13]; and rapid science will be critical for progress [14].

## Data Availability

Data sources, statistical code, and supporting documents are available here: https://github.com/andrewlover/HCQ_AZ_COVID_19

## Revision History

| Revision | Date | Author(s) | Description |
| --- | --- | --- | --- |
| 1.0 | 22.3.20 | AAL | created |
| 1.1 | 23.3.20 | AAL | updated survival models; minor edits |
| 1.2 | 24.3.20 | AAL | minor edits |
| 1.3 | 04.1.20 | AAL | addition of preface statement and associated references |

